# SalivaAll: Clinical validation of a sensitive test for saliva collected in healthcare and community settings with pooling utility for SARS-CoV-2 mass surveillance

**DOI:** 10.1101/2020.08.26.20182816

**Authors:** Nikhil S Sahajpal, Ashis K Mondal, Sudha Ananth, Allan Njau, Pankaj Ahluwalia, Alka Chaubey, Vamsi Kota, Kevin Caspary, Ted M Ross, Michael Farrell, Michael P. Shannon, Amyn M Rojiani, Ravindra Kolhe

## Abstract

**Background:** The adoption of saliva as a specimen type for SARS-CoV-2 mass surveillance can significantly increase population compliance with decreased exposure risk for healthcare workers. However, studies evaluating the clinical performance of saliva compared to nasopharyngeal swab (NPS) samples have demonstrated conflicting results regardless of the collection being in healthcare or community settings. Further, pooled testing with saliva remains a challenge owing to the ambiguous sensitivity, limit of detection (LoD), and processing challenges. To overcome these limitations, SalivaAll protocol was developed and validated as a cost-effective measure that must be used on saliva collected in health care or community settings with pooling utility for SARS-CoV-2 mass surveillance.

**Methods:** The study evaluated 429 matched NPS and saliva samples collected from 344 individuals in either healthcare or community setting. In phase I (protocol U), 240 matched NPS, and saliva samples were tested for SARS-CoV-2 detection by RT-PCR. In phase II (SalivaAll protocol), 189 matched NPS and saliva samples were tested, with an additional sample homogenization step for saliva before RNA extraction, followed by RT-PCR. Eighty-five saliva samples were evaluated with both protocols (U and SalivaAll). Subsequently, adopting SalivaAll protocol, a five-sample pooling strategy was evaluated for saliva samples based on FDA recommendations.

**Results:** In phase I, 28.3% (68/240) samples tested positive for SARS-CoV-2 from either saliva, NPS, or both. The detection rate was lower in saliva compared to NPS samples (50.0% vs. 89.7%). In phase II, 50.2% (95/189) samples tested positive for SARS-CoV-2 from either saliva, NPS, or both. The detection rate for SARS-CoV-2 was higher in saliva compared to NPS testing (97.8% vs. 78.9%). Of the 85 saliva samples evaluated by both protocols, 57.6% (49) tested positive for SARS-CoV-2 with either protocol U, SalivaAll, or both. The detection rate was 100% for samples tested with SalivaAll, whereas it was 36.7% with protocol U. Also, the LoD with SalivaAll protocol was 20 copies/ml. The pooled testing approach demonstrated a 95% positive and 100% negative percent agreement.

**Conclusion:** This single-site study demonstrated the variability of results reported in the literature for saliva samples, and found that the discrepancies are explained by processing challenges associated with saliva samples. We have optimized a protocol for saliva samples that results in higher sensitivity compared to NPS samples and also breaks the barrier to using pooled saliva testing for SARS-CoV-2.

**Summary:** SalivaAll is a very sensitive (LoD 20 copies/ml) cost-effective test validated on saliva collected in health care and community settings with pooling utility and submitted for FDA Emergency Use Authorization.

## Introduction

The outbreak of COVID-19 (caused by SARS-CoV-2), is an ongoing pandemic that has caused substantial social, economic, and public health strain. Since its identification in the region of Wuhan, China, 22,847,562 confirmed cases with over 797,248 COVID-19 related deaths have been reported globally (https://coronavirus.jhu.edu/map.html, last accessed August 21, 2020). Testing, coupled with measures such as social distancing, masking, maintaining personal hygiene, contact tracing, quarantine, travel restrictions, and lockdowns have hitherto been the most widely adopted strategies to contain this pandemic. [1]. Regional policies have been primarily dictated by the positivity rate in the respective region(s). Thus, rapid and accurate detection of SARS-CoV-2 is the foremost and likely most essential component in controlling this outbreak and has immediate clinical, epidemiological, and policy implications. The detection of the SARS-CoV-2 virus has been applied to various clinical specimens that include bronchoalveolar lavage, sputum, saliva, nasopharyngeal swabs (NPS), oropharyngeal swabs (OPS), feces, and blood [2]. NPS and OPS samples are the current standard upper respiratory tract specimens recommended for COVID-19 diagnostic testing. However, the collection of NPS samples poses certain challenges that include exposure risk to healthcare workers, supply chain constraints pertaining to swabs and personal protective equipment, and self-collection being difficult and less sensitive. Furthermore, several reports have highlighted the relatively poor sensitivity of NPS samples in early infection and longitudinal testing [3-5]. Amidst these challenges, several other sample types have been under investigation for COVID-19 testing, of which, saliva samples are of significant interest owing to their ease of collection, and alleviating some of the challenges associated with NPS sampling. “True” saliva is defined as the naturally collecting clear liquid that accumulates in the mouth. However, saliva from patients can be confounded with the presence of mucus or blood, thereby rendering it a difficult sample to process in the laboratory. Several reports evaluating the clinical performance of saliva compared to NPS/OPS samples have demonstrated conflicting results. In a healthcare setting, studies have demonstrated comparable [6-9] and even higher sensitivity of saliva [10]/ early morning saliva collection [11] compared to NPS samples, as well as reported higher viral titer values in saliva [9]. Conversely, deep-throat saliva [12] and typical saliva samples have also been demonstrated to be less sensitive compared to NPS samples in both healthcare [13] and community settings [14]. Furthermore, saliva samples are difficult to pipet by the testing personnel, which leads to increased processing time [15].

Although different collection devices, media, sample handling, extraction procedure, and RT-PCR methods may have accounted for these discrepancies; we investigated a critical facet of the saliva samples that if accounted for, renders the saliva samples more sensitive than NPS samples. In our analysis, the saliva samples initially demonstrated a lower sensitivity compared to NPS samples. We introduced a simple processing step using the bead mill homogenizer and observed that the saliva samples showed higher sensitivity compared to NPS samples. Not only did the saliva processing became significantly easier after using the homogenizer, but we were also able to validate saliva samples with a five-sample pooling strategy. The SalivaAll protocol is very sensitive and detects up to 20 copies/ml, facilitating utility in saliva sample pooled testing which will be critical in SARS-CoV-2 mass surveillance. Our single-site study identified both increased and decreased sensitivity of saliva as a diagnostic sample type, consistent with the discrepancies reported in the literature, that were largely believed to be the result of processing challenges. We contend that with appropriate management of these processing challenges saliva samples are more sensitive compared to the NPS samples.

### Material and Methods

### Study site and ethics

This single-center diagnostic study was conducted at Augusta University, GA, USA. This site is a CLIA accredited laboratory for high complexity testing and is one of the main SAR-CoV-2 testing centers in the state of Georgia.

### Patient Specimens and setting

The study evaluated 429 matched NPS and saliva samples collected from 344 individuals in either a healthcare or community setting. Of the 344 individuals, 95 matched clinical specimens were collected in healthcare setting from individuals either at a Medical nursing home, GA, or Augusta University Medical Center, GA. In the community setting, 249 matched clinical specimens were collected from drive-through collection centers in different regions of Georgia that include Augusta, Albany, and Atlanta. As a standard protocol in both settings, NPS from individuals was collected by a healthcare worker using a sterile flocked swab placed in a sterile tube containing the Viral transport medium (VTM). Before collecting the NPS samples, the individuals were instructed to provide saliva samples by spitting into a sterile container over which the healthcare worker added the VTM. All samples were stored at 4°C temperature and transported to the SARS-CoV-2 testing facility at Augusta University, GA within 24 hours of sample collection, for further processing.

### Assay for the detection of SARS-CoV-2

The assay is based on nucleic acid extraction followed by TaqMan-based RT-PCR assay to conduct in vitro transcription of SARS-CoV-2 RNA, DNA amplification, and fluorescence detection (FDA-EUA assay by PerkinElmer Inc. Waltham, USA). The assay targets specific genomic regions of SARS-CoV-2: nucleocapsid (N) gene and *ORF1ab*. The TaqMan probes for the two amplicons are labeled with FAM and ROX fluorescent dyes, respectively, to generate target-specific signals. The assay includes an RNA internal control (IC, bacteriophage MS2) to monitor the processes from nucleic acid extraction to fluorescence detection. The IC probe is labeled with VIC fluorescent dye to differentiate its fluorescent signal from SARS-CoV-2 targets.

### Phase I study and sample processing (Protocol U)

In phase I of this study, 240 matched NPS and saliva samples were tested prospectively for SARS-CoV-2 RNA by RT-PCR. Of the 240 samples, 95 samples were collected in a healthcare setting and 145 were collected in a community setting. In brief, an aliquot of 300μl from each sample (NPS or Saliva), positive and negative controls, were added to respective wells in a 96 well plate. To each well, 5μl internal control (IC), 4μl Poly(A) RNA, 10μl proteinase K and 300μl lysis buffer 1 were added. The plate was placed on a semi-automated instrument (chemagic 360 instrument, PerkinElmer Inc.) following the manufacturer’s protocol. The nucleic acid was extracted in a 96 well plate, with an elution volume of 60μl. From the extraction plate, 10μl of extracted nucleic acid and 5μl of PCR master mix was added to the respective wells in a 96 well PCR plate. The PCR method was set up as per the manufacturer’s protocol on Quantstudio 3 or 5 (ThermoFisher Scientific, USA). The samples were resulted as positive or negative, based on the Ct values specified by the manufacturer (**Figure 1**).

**Figure 1.**
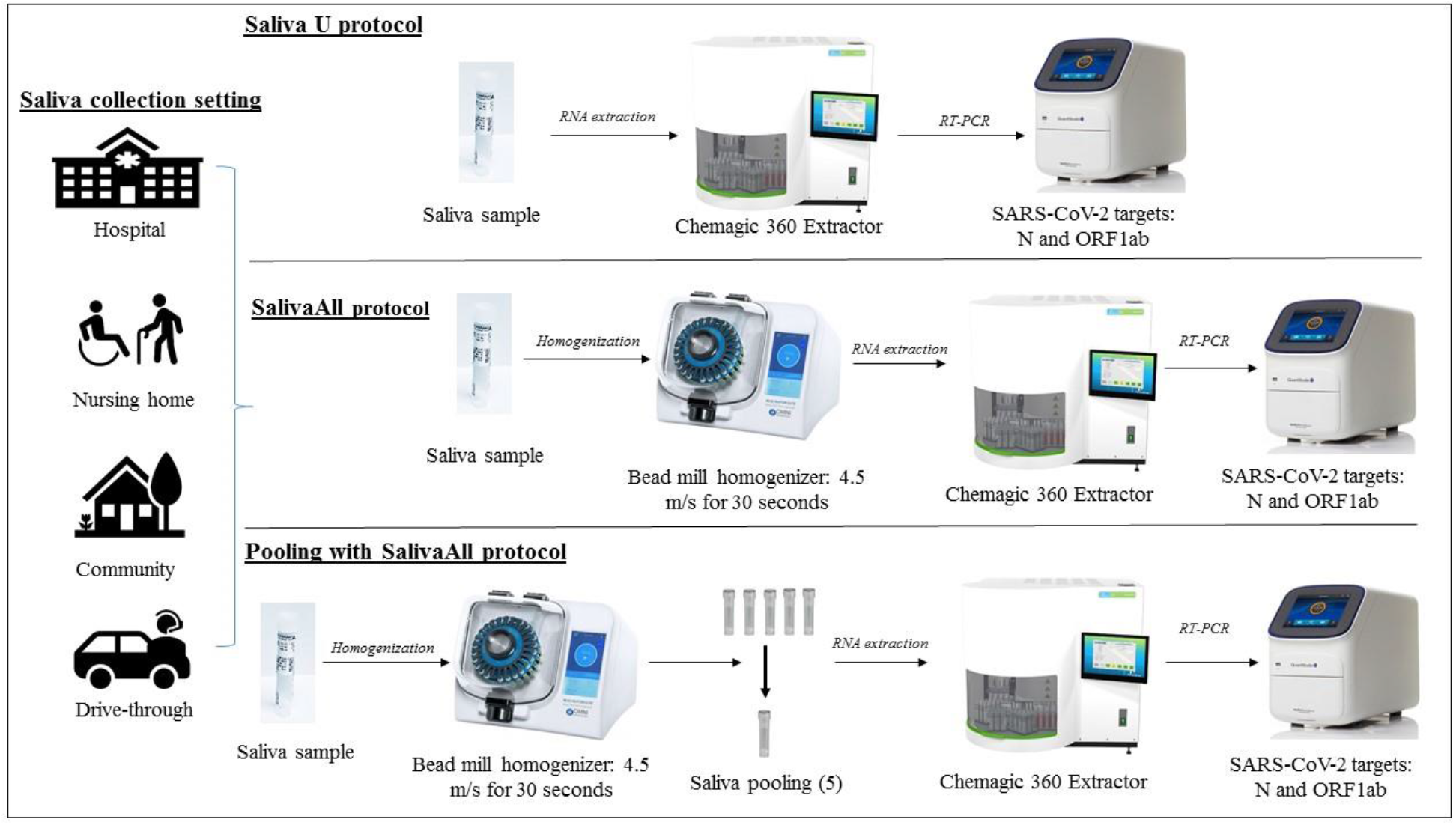
Schematic overview of sample processing and SARAS-CoV-2 assay workflow depicting main steps: Matched nasopharyngeal swabs and saliva samples collected in healthcare and community setting were tested and validated as follows: Upper panel: NPS or saliva samples were processed with protocol U for Nucleic acid extraction using a semi-automated instrument, followed by RT-PCR for N, ORF1ab gene targets and IC used as extraction and RT-PCR internal control; Middle panel: Saliva samples processed with SalivaAll protocol that included a saliva homogenization step using a bead mill homogenizer before RNA extraction and downstream processing; Lower panel: Saliva samples were homogenized using a bead mill homogenizer (SalivaAll protocol) before pooling samples with a five-sample pooling strategy for SARS-CoV-2 testing.

### Phase II study and sample processing (SalivaAll)

In phase II of this study, 189 matched NPS and saliva samples were tested for SARS-CoV-2. Of the 189 samples, 40 were collected in healthcare, and 149 were collected in a community setting. More importantly, 85 samples that had been previously evaluated with protocol U were reevaluated with SalivaAll. In SalivaAll protocol, an additional processing step was added for saliva samples that included aliquoting saliva samples from the collection tubes into Omni tubes, which were then placed in Omni bead mill homogenizer (Omni International, USA). The samples were homogenized at 4.5 m/s for 30 seconds. From the Omni tubes, an aliquot of 300μ1 homogenized saliva samples were added to 96 well plate and downstream processing was carried out as described in protocol U. The NPS sample processing remained the same as in protocol U (**Figure 1**).

### Limit of detection studies

The limit of detection (LoD) studies were conducted as per the FDA guidelines (https://www.fda.gov/medical-devices/coronavirus-disease-2019-covid-19-emergency-use-authorizations-medical-devices/vitro-diagnostics-euas). Briefly, SARS-CoV-2 reference control material (PerkinElmer Inc.) was spiked into the negative saliva samples to serve as positive samples at 180 copies/ml, 60 copies/ml and 20 copies/ml concentrations, and were processed with SalivaAll protocol. The lowest concentration detected in all three triplicates was determined as the preliminary LoD. To confirm the LoD, 20 replicates of preliminary LoD were analyzed and deemed as confirmed if at least 19/20 replicates were detected.

### Pooling saliva samples for Mass Population screening

A five-sample pooling strategy was evaluated as per FDA guidelines (https://www.fda.gov/medical-devices/coronavirus-disease-2019-covid-19-emergency-use-authorizations-medical-devices/vitro-diagnostics-euas). Briefly, 20 previously confirmed positive saliva samples were identified to create 20 positive pools each comprising of one positive and four negative samples. The Ct values of positive samples ranged from (N: 9.8, *ORF1ab:* 17.3 - N: 38.4, *ORF1ab:* Undetermined), with 25% of positive samples having high Ct values (32.0 - 38.4). Similarly, 20 negative sample pools were created comprising of five negative samples. All saliva samples were first processed with Omni bead mill homogenizer as described in SalivaAll protocol before pooling for SARS-CoV-2 RT-PCR testing (**Figure 1**). Of note, each pool was created by aliquoting 60μl from each of the five homogenized samples to make 300μl for extraction.

## Results

### Comparison of SARS-CoV-2 detection between saliva and NPS (Phase I study - Protocol U)

Of the 240 participants, 28.3% (68/240) tested positive for SARS-CoV-2 from either saliva, NPS, or both. The detection rate for SARS-CoV-2 was higher in NPS compared to saliva testing (89.7% [61/68] vs. 50.0% [34/68]). The concordance for positive results between the two tests was only 39.7% (virus detected in both saliva and NPS in 27/68). Of the 68 positive samples, 50% (34/68) of samples resulted positive in NPS but not in saliva, while only 10.2% (7/68) resulted positive in saliva but not in NPS. The Ct values for *N*(25.2 ± 8.7 vs. 29.5 ± 6.4, p<0.05), *ORF1ab* (22.6 ± 7.9 vs. 33.5 ± 6.0, p < 0.001) and IC (32.2 ± 3.5 vs. 36.0 ± 4.4, p<0.001) were significantly lower in NPS compared to saliva samples, respectively **(Figure 2a and b)**. Overall, IC (extraction and RT-PCR control) was detected in 95% (228/240) NPS samples, and 80.4% (193/240) saliva samples. **Comparison of SARS-CoV-2 detection between saliva and NPS (Phase II study - SalivaAll)** Of the 189 samples tested in Phase II, 50.2% (95/189) tested positive for SARS-CoV-2 from either saliva, NPS, or both. The detection rate for SARS-CoV-2 was higher in saliva compared to NPS testing (97.8% [93/95] vs. 78.9% [75/95]). The concordance for positive results between the two tests was 76.8% (virus detected in both saliva and NPS in 73/95). Of the 95 positive samples, 21.0% (20/95) of samples resulted positive in saliva but not in NPS, while only 2.1% (2/95) resulted positive in NPS but not in saliva. The Ct values for *N* (26.9 ± 8.0 vs. 26.4 ± 6.9), *ORF1ab* (27.0 ± 8.6 vs. 29.2 ± 6.3) were comparable, whereas the IC values (33.4 ± 3.6 vs. 31.6 ± 3.4, p<0.05) were significantly higher in NPS compared to saliva samples, respectively **(Figure 2c and d)**. Overall, IC (extraction and RT-PCR control) was detected in all except one saliva sample.

**Figure 2:**
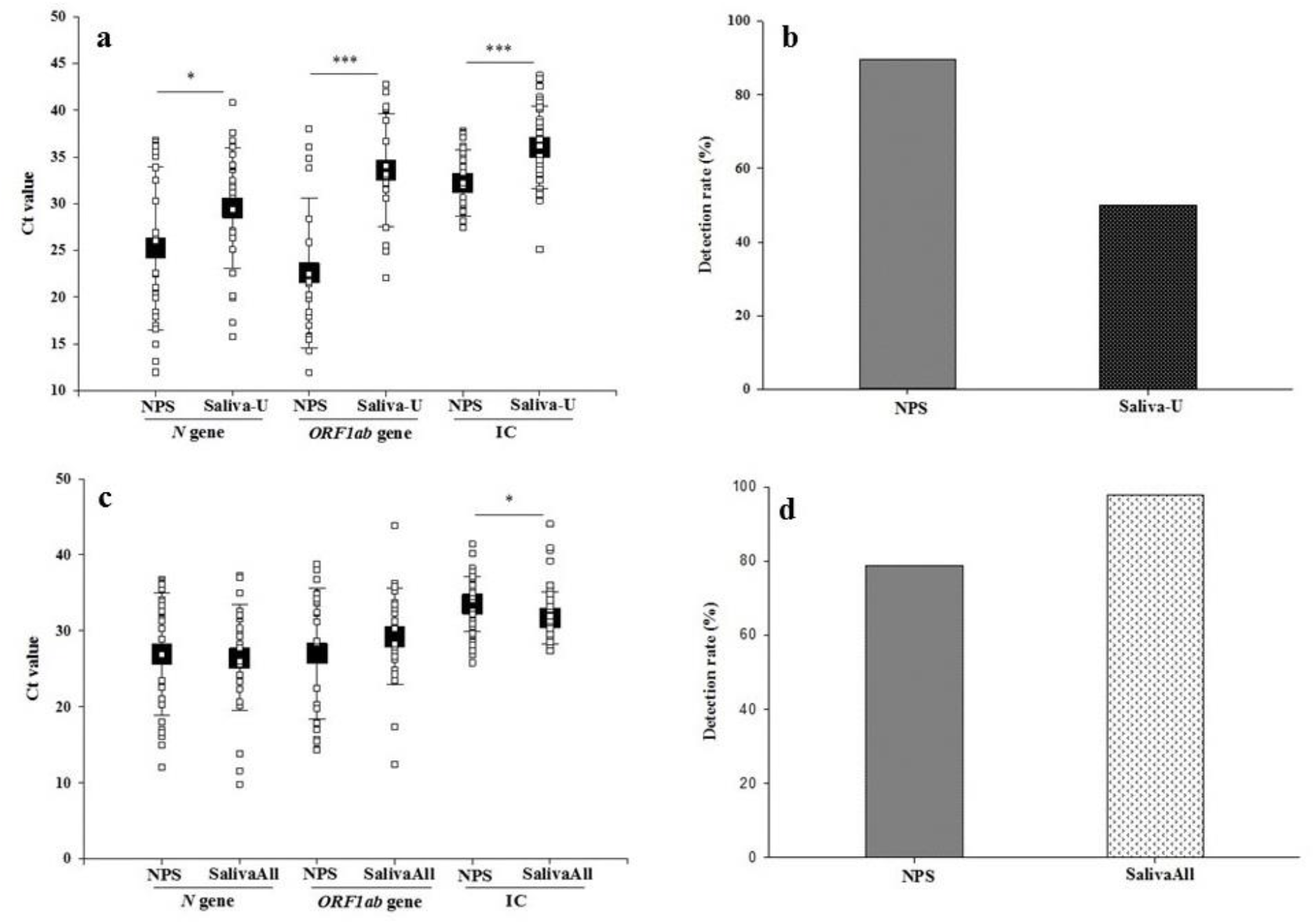
**a**. Boxplots of SARS-CoV-2 Ct value (mean±SD) of both N gene and ORF1ab-gene of all the positive specimens in phase I study. The Ct value of both genes was lower in the NPS than Saliva samples. Also included Ct values of internal control (IC) of all the samples in both specimens. **b**. Bar graph depicting detection rate with NPS and saliva samples in phase I study.**c**. Boxplots of SARS-CoV-2 Ct value (mean±SD) of both N gene and ORF1ab-gene of all the positive specimens in phase II study. The Ct value of both genes was lower in the saliva than NPS samples. Also included Ct values of internal control (IC) of all the samples in both specimens. **d**. Bar graph depicting detection rate with NPS and saliva samples in phase II study

### Comparison of SARS-CoV-2 detection in saliva samples (Protocol U vs. SalivaAll)

Of the 85 saliva samples, 57.6% (49/85) tested positive for SARS-CoV-2 with either protocol U, SalivaAll, or both. The detection rate was 100% (49/49) with samples tested with SalivaAll, whereas it was only 36.7% (18/49) with protocol U. The concordance for positive results between the two protocols was 36.7% (18/49). The Ct values for *N* (25.1 ± 6.0 vs. 28.6 ± 5.9) was found to be lower whereas, *ORF1ab* (27.9 ± 6.0 vs. 35.2 ± 6.3, p < 0.01) and IC (31.2 ± 2.7 vs. 33.7 ± 3.9, p<0.001) were significantly lower with SalivaAll compared to protocol U, respectively **(Figure 3a and b)**.

**Figure 3:**
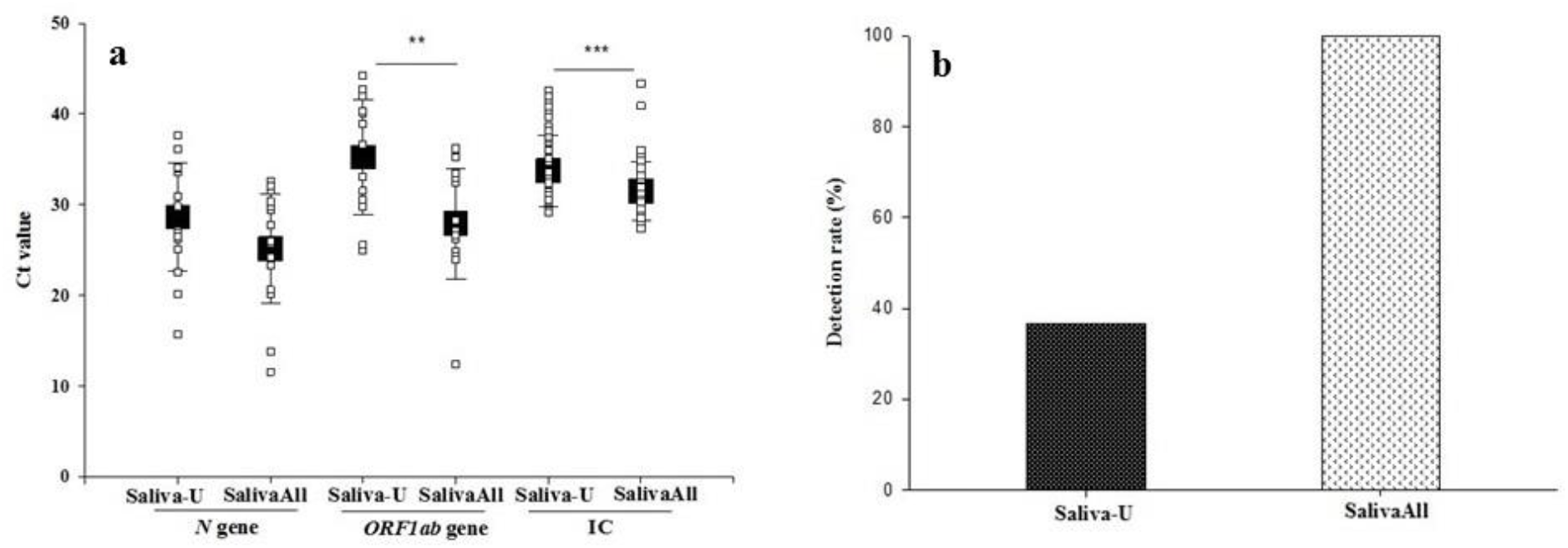
**a**. Boxplots of SARS-CoV-2 Ct value (mean±SD) of both N gene and ORF1ab-gene of all the positive saliva specimens detected with protocol U and B. Ct value of both genes were lower SalivaAll than A. Also included Ct values of internal control (IC) of all the samples in both specimens. **b**. Bar graph depicting the detection rate in the saliva sample with protocol U and SalivaAll.

### Limit of detection

In the preliminary LoD study, all replicates were detected at the three tested concentrations. The LoD was determined to be 20 copies/ml, and all 20 replicates were detected (**Table 1a**).

**Table 1a.** SARS-CoV-2 limit of detection using SalivaAll protocol.

| SARS-CoV-2<br>(triplicates) | N gene<br>(Mean Ct ± SD) | ORF1ab gene<br>(Mean Ct ± SD) |
| --- | --- | --- |
| 20 copies/ml | 36.7 ± 0.6 | 35.1 ± 1.3 |
| 60 copies/ml | 33.6 ± 0.15 | 33.7 ± 0.4 |
| 180 copies/ml | 32.7 ± 0.19 | 32.6 ± 0.2 |
| **LoD confirmation (20 copies/ml) : 20/20 replicates detected** |  |  |

### Pooling saliva samples for Mass Population screening

The five-sample pooling strategy was evaluated by comparing the results of the 20 positive and negative pools to individual sample testing results. The pooled testing results demonstrated a 95% positive and 100% negative percent agreement. The *N* and *ORF1ab* gene Ct values were compared between pooled and individual testing. Regression analysis with slope and intercept along with a 95% confidence interval was determined. The shift in Ct value was found to be significant with pooled testing towards higher Ct values, nonetheless, the pools containing positive samples with viral loads close to the assay’s LoD (i.e., weak positives) were accurately detected (**Table 1b**) (**Figure 4a and b**).

**Table 1b.** Performance of saliva pooling with SalivaAll protocol.

| Study design-SalivaAll | Composition of Saliva pools | Concordance |
| --- | --- | --- |
| **Positive pools (20)** | 1 positive + 4 negative (Individually tested samples) | 19/20 |
| **Negative pools (20)** | 5 negative (Individually tested samples) | 20/20 |
| **Positive percent agreement = 95%** |  |  |
| **Negative percent agreement = 100%** |  |  |

**Figure 4:**
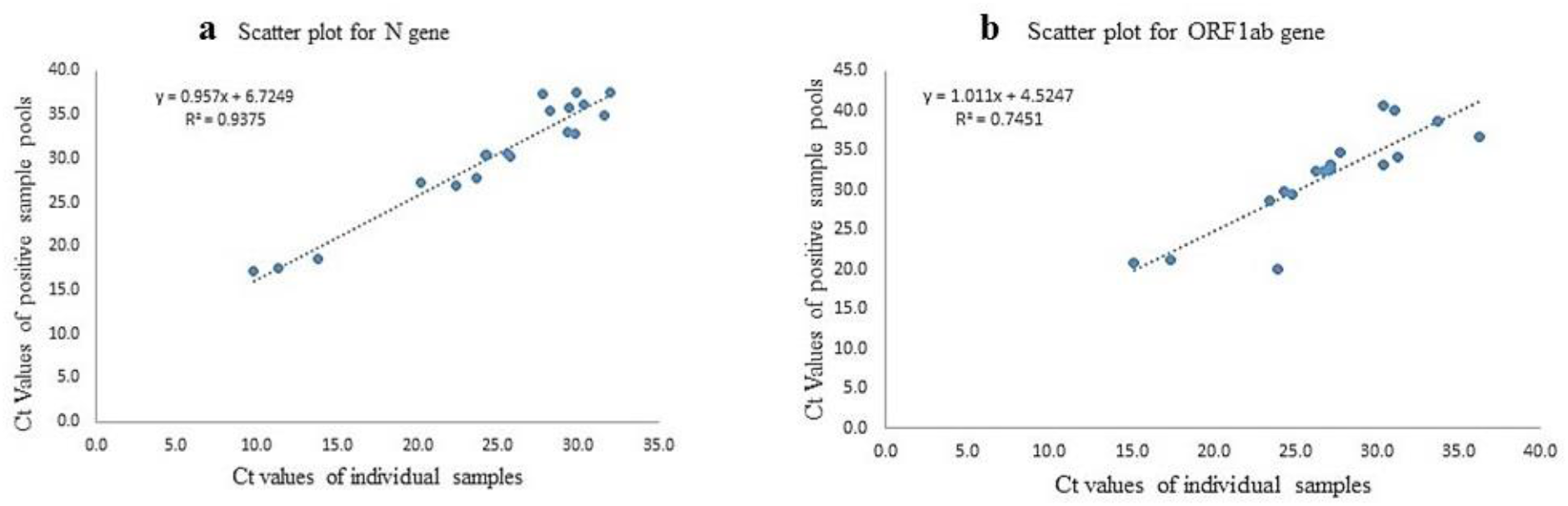
**a**. The Ct values comparison of N gene with individual testing vs. pool testing. **b**. The Ct values comparison of ORF1ab gene with individual testing vs. pool testing.

## Discussion

An optimal sample type and easier collection method for the detection of SARS-CoV-2 are primary requirements in the global effort to control this pandemic. NPS and OPS samples are the currently recommended sample types for COVID-19 diagnostic testing. Testing for SARS-CoV-2 has relied primarily on these samples, but the associated challenges with NPS/OPS sampling have engendered a need to evaluate several other sample types, of which saliva has remained an attractive alternate. However, there are conflicting reports in the literature on the clinical performance of saliva compared to NPS/OPS samples. In a healthcare setting, initial reports from To et al. demonstrated comparable sensitivity of saliva samples (86.9% and 91.7%) compared to NPS samples [6,7]. Similarly, independent investigations from Azzi et al. and Yoon et al., demonstrated a 100% concordance of saliva with NPS samples, with Yoon et al., reporting high viral titer values in saliva [8,9]. In addition, Wylie et al. and Rao et al. demonstrated higher sensitivity of saliva and particularly early morning saliva compared to NPS samples, respectively [10,11]. Conversely, Lai et al. and Jamal et al. reported lower sensitivity of deep-throat saliva (68.7% vs. 80.9%) and saliva samples (72% vs. 89%) compared to NPS samples, respectively [12,13]. Furthermore, Becker et al. reported a 30-50% lower sensitivity of saliva compared to NPS samples in the community setting [14].

In phase I of our study comprising 240 matched NPS and saliva samples (protocol U), we observed lower sensitivity of saliva samples compared to NPS samples. The detection rate in saliva samples was significantly lower compared to NPS samples. In addition, the Ct values for N, *ORF1ab* and IC (extraction and PCR control) were significantly higher in saliva compared to the NPS sample. Further, a high percentage of saliva samples yielded invalid results. A significant challenge in the wet lab revolved around accurate pipetting of saliva samples since the majority of these were very viscous. The viscous gel-like consistency not only caused issues with pipetting but also led to a high percentage of invalid results. This was demonstrated as a significantly high Ct value drift in the internal control (IC) suggesting that the lower sensitivity of saliva samples is most likely due to processing and sometimes pre-analytical issue(s). Several factors that might have led to the lower sensitivity of saliva samples were contemplated including inaccurate pipetting of sample volume into the extraction plate for nucleic acid extraction, the viscous gel-like texture of saliva samples prevented appropriate mixing with viral transport media which led to the pipetting of primarily “saliva” or “media” into extraction plate for nucleic acid extraction, the likelihood that the higher viscosity of saliva might also impair sample lysis for nucleic acid extraction.

The underlying issue associated with these challenges emerged to be the gel-like consistency of saliva samples. We addressed this significant preanalytic concern by adding a simple step before processing samples for nucleic acid extraction. The saliva samples were homogenized using a bead mill homogenizer at a speed of 4.5 m/s for 30 seconds (SalivaAll). The homogenization rendered the saliva samples to uniform viscosity and consistency, making it easier to pipet for the downstream assay. Thus, in phase II of the study with 189 matched NPS and saliva samples, adding this single pre-processing step rendered saliva samples more sensitive compared to NPS samples. The detection rate was higher in saliva samples, with significantly lower Ct value for IC, compared to NPS samples. Also, on comparing the 85 saliva samples that were processed both in phase I and phase II studies, the detection rate was significantly higher in samples processed with the homogenization step (SalivaAll) compared to the sample processed without it (protocol U). The Ct values in the same saliva samples were lower for the *N* gene, and significantly lower for *ORF1ab* and IC with samples processed with SalivaAll compared to protocol U. These results demonstrate that the lower sensitivity of the saliva samples observed in the phase I study was because of inadequate sample processing. The saliva samples were found to be more sensitive compared to NPS samples, with the addition of this simple processing step. Further, the LoD of 20 copies/ml confirmed the sensitivity of saliva samples as the LoD was determined to be the same as that for the FDA-EUA comparator test for NPS samples.

Furthermore, saliva processing became significantly easier, and we were also able to successfully validate saliva samples with a five-sample pooling strategy. The pooled testing results demonstrated a positive percent agreement of 95% (19/20 pools showing positive results), with one pool that contained the sample with very high Ct (N: 38.4; ORF1ab: undetermined) being undetectable. The negative percent agreement was found to be 100%.

We have previously demonstrated the feasibility and accuracy of a sample pooling approach with NPS samples for wide-scale population screening for COVID-19 [16]. Herein, we extend the utility and potential benefits of the sample pooling approach for population screening with saliva samples. Considering the evolving epidemiology of COVID-19 and the reopening of educational and professional institutions, travel, tourism, and social activities, monitoring SARS-CoV-2 will remain a critical public health need. Therefore, the use of a non-invasive collection method and easily accessible sample such as saliva will enhance screening and surveillance activities.

In conclusion, this study has demonstrated that the saliva samples are more sensitive than NPS samples if collected and processed appropriately before extraction and PCR. The study evaluated matched NPS and saliva samples collected in both healthcare and community settings. In the community setting, no control was exercised regarding food or drink restriction, and time of sample collection as most of the community-collected samples came from drive-through facilities that operates from morning until evening. The spectrum of disease also varied from asymptomatic to severely ill patients, and the study was not biased for a particular group. However, the study has two limitations that the Ct-value in this study reflects viral load but not the viral copies per ml, and the sample collection were not controlled for a specific day of illness. Nonetheless, despite these limitations, this study presents a significant and clinically validated approach to the utilization of saliva samples for Covid-19 testing, individually or as pooled samples.

## Data Availability

All data is available within the manuscript.

## Acknowledgment

The authors would like to thank Dr. Brooks Keel, President, Augusta University and AUHS for his strategic, science-based, data-driven leadership in all matters COVID-19, and constant support and encouragement at every step of this work.

## Notes

### Competing Interest Statement

The authors have declared no competing interest.

### Funding Statement

No funding was received for this work.

### Author Declarations

All studies were performed under a approved protocol (HAC 611298) by the Institutional Review Board at Augusta University.

